# A shred of evidence that BCG vaccine may protect against COVID-19: Comparing cohorts in Spain and Italy

**DOI:** 10.1101/2020.06.05.20123539

**Authors:** David I. Levine

**Affiliations:** University of California, Berkeley

## Abstract

**Introduction:** There is evidence that the BCG vaccine against tuberculosis also helps prevent other diseases – perhaps including COVD-19. Spain had a program for universal BCG vaccination until 1981.

**Objective:** To see whether cohorts born when Spain had a program of universal BCG vaccination had lower rates of confirmed cases of COVID-19 and mortality (relative to similar cohorts in Italy).

**Methods:** We compare COVID-19 mortality and confirmed cases for those born roughly a decade before and after 1981. We compare the outcomes to the same age cohorts in Italy, which never had universal BCG vaccination.

**Results:** The Spanish cohort that received BCG had a relative risk of 0.962 of having a confirmed case of COVID-19. This risk is statistically significantly below unity (95% CI 0.952 to 0.972, P< 0.001). There is also suggestive evidence the BCG cohort in Spain had lower mortality (relative risk 0.929, CI 0.850 to 1.01, P = 0.11). The small sample size makes this test underpowered.

**Conclusion:** These suggestive results provide a shred of evidence that BCG vaccinations help protect against COVID. I outline many limitations to this study and point how better data can help be more convincing.

---

There is evidence that the BCG vaccine against tuberculosis strengthens the immune system and prevents diseases beyond tuberculosis.^2^ Perhaps coincidentally, nations that never had universal BCG vaccinations (such as the United States and Italy^3^) have higher rates of reported COVID-19 cases than nations that had or have widespread BCG vaccination.^4^ These results have motivated two randomized trials of whether BCG prevents COVID among health workers.^5^

At the same time, these cross-national data are not very convincing because nations with ongoing BCG vaccination typically are poor nations with lower rates of testing for COVID-19. ^6^ Most nations with BCG are also less connected to China, so may just be slower to start moving up the COVID curve.

We can extend the cross-national studies by examining data from Spain, a nation that had a program for universal BCG vaccination from 1965 to 1981. We look at those born just before and just after Spain ended its BCG program. To adjust for the effects of age on COVID outcomes, we compare COVID confirmed cases and mortality in Spain to those in Italy. Italy resembles Spain in many ways, but never had a program for universal BCG. Our objective is to see whether cohorts born when Spain had a program of universal BCG vaccination had lower rates of confirmed cases of COVID-19 and mortality (relative to similar cohorts in Italy).

## Methods

### The setting

We briefly review the two nations we study, COVID-19, and the BCG vaccine.

**Spain and Italy** are nearby nations with many similarities. Prior to COVID they had similar life expectancies (82.8 in Spain, 82.7 in Italy), both among the highest in Europe. Both also have high-quality health systems that are largely free of cost and run by the public sector. Spain and Italy resemble each other on a host of measures such as age distribution, rates of obesity, rates of diabetes, and hospital beds per 1000 population (Table 1).

**Table 1:** Indicators in Spain and Italy

|  | Spain | Italy | Global |
| --- | --- | --- | --- |
| Obesity | 21% | 24.7% | 13% |
| Diabetes (ages 20-79) | 6.9% | 5.0% | 8.8% |
| Age > 65 | 19% | 23% | 8.9% |
| Cause of death, by communicable diseases and maternal, prenatal and nutrition conditions (% of total) | 5% | 5% | 20.2% |
| Hospital beds (per 1000 population) | 3.0 | 3.4 | 2.7 |

Obesity data are from https://www.worldobesitydata.org/map/overview-adults (Italy and Spain) and from WHO "Obesity and Overweight." April 1, 20202. https://www.who.int/news-room/fact-sheets/detail/obesity-and-overweight. Other data are from World Bank Indicators. https://data.worldbank.org/indicator [Last accessed May 13, 2020]

**COVID-19** was first recognized in December 2019 in Wuhan China. COVID-19 is a serious respiratory infection caused by the coronavirus SARS-COV2. It has a high mortality rate among confirmed cases. Mortality is strongly associated with advanced age and with comorbidities such as cancer, COPD, hypertension, and diabetes.^7^

COVID-19 quickly spread around the globe. Community transmission began in both Italy and Spain in mid-February 2020.^8^ By the end of March 2020, the two nations had the most confirmed cases in the world.^9^ A month later, each nation had hundreds of thousands of confirmed cases and tens of thousands of deaths. At the time of our data collection, both nations had similar rates of mortality attributed to COVID and of COVID testing per million population.^10^

**BCG (Bacillus Calmette-Guérin)** is the most common vaccine for tuberculosis (TB). At the same time, as noted above, several studies find that BCG lowers the risk of many other diseases.^11^ This evidence suggests that BCG strengthens the fast-acting innate immune system.^12^

Spain had a nation-wide program for universal BCG vaccination at birth from 1965 to 1981.^13^

In contrast, Italy has never had a universal program for BCG. Both nations always immunized specific high-risk populations with the BCG vaccine.

### Data

We examine two outcomes: mortality reported to be due to COVID-19 and confirmed cases of COVID-19. We rely on published data on for 10-year age groupings in each nation. The data for Spain are from the Ministerio de Sanidad, Consumo y Bienestar Social and cover until 1 May 2020. ^14^ The data for Italy are from Instituto Superiore di Sanità and cover until 28 April 2020.^15^

Both outcomes are subject to substantial under-reporting.

First, many cases of COVID-19 are never confirmed. Both Italy and Spain had shortages of tests during the first months of the epidemic - the period we study.^16^ In addition, many cases are asymptomatic or nearly asymptomatic.^17^ Thus, the rates of *confirmed* cases will miss many cases, especially those with milder symptoms. In addition, the mortality data are incomplete as some victims of COVID-19 die without a confirmed diagnosis. That “missing mortality” is concentrated among those who die in nursing homes. We study a population under age 50, so we expect fewer missing deaths.

Fortunately, unless the rates of under-reporting differed across the two cohorts *and* the differential across cohorts differs in the two nations, the under-reporting of COVID-19 cases or mortality will not bias our tests. In fact, the more severe symptoms that receive a confirmed diagnosis may be an outcome of greater interest than examining all COVID-19 cases.

We also use data on the population distribution by age and sex from https://Populationpyramid.net. These data are of the form: “Men ages 30–34 are 2.9% of the Spanish population,” which introduces rounding error.

As others have found, age has a very strong effect on mortality from COVID-19. Thus, only about 1% of the deaths in Italy and Spain are ages 30 to 49.

In both nations, mortality is higher is higher for men than for women. In contrast, women have more confirmed diagnoses than men.

### Statistical methods

Recall that Spain stopped universal BCG in 1981. During the study period (February to April 28 or May 1, 2020), almost everyone born before 1981 was 40 or older. Conversely, everyone born after 1981 is 39 or younger. The core analysis is to compare outcomes among those in their 30s versus those in their 40s in each nation. If BCG is protective against COVID, then the relative risk of people in their forties should be lower in Spain. That is, for each outcome *Y*, where *Y* = mortality attributed to COVID-19 or a confirmed COVID-19 case, we test whether the relative risk

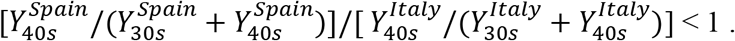

Although this hypothesis is one sided, the preliminary nature of this analysis leads me to use a two-sided test of statistical significance. Also limited sample size for mortality leaves that analysis underpowered. Thus, it is important to give low emphasis to those results.

### Adjusting for differences in the age distributions

Spain and Italy do not have the same distribution of ages within each decade. With some assumptions, we can estimate what the Spanish outcomes would have been if Spain had had the Italian age distribution.

First, we take the outcome counts from the health data and the population counts (from national data) to estimate outcome ratios in each nation (*n*) for each decade cohort (*d*), *R_nd_*; for example, confirmed cases per capita for people in their thirties in Spain. “Ratios” here refer to shares of the national population of that age (or age-and-sex) cohort.

We assume the growth rate in outcome ratios decade-to-decade in Italy is the same for each half-decade in each nation. Thus, the growth rate in each half-decade is:

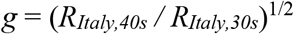

For example, Italy has 1.33 times as many confirmed cases per capita in their 40s than in their 30s. So this growth rate across decades is consistent with 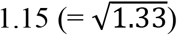 more cases per half decade.

Let *S_nd_* = the share of each decade’s cohort in each nation in the younger half of that decade.

We then apply that within-decade growth rate to each nation so that, coupled with the population share, would yield the observed average outcome ratio in that nation for each half-decade cohort. These assumptions imply the outcome ratio (*Ŷ*) for the younger 5-year subcohort of each decade (subscripted *d,young*) is:

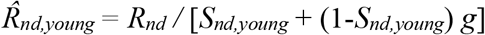

The predicted outcome ratio for the older half of each decade is:

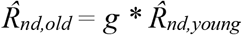

Finally, we weight the Spanish predicted outcome ratios by half-decade by the *Italian* population shares to estimate a counterfactual outcome ratio by half-decade that Spain would have experienced if they had had the Italian age distribution:

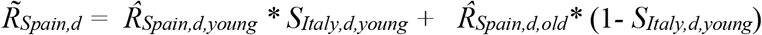

This procedure is reasonable give the available data, but it is also very rough.

## Results

In Italy there are 1.70 confirmed cases among people in their 40s for each case among those in their 30s (31,488 vs. 20,282). In Spain, in contrast, there are about 1.55 confirmed cases of people in their 40s for each case among those in their 30s (25,644 vs. 14,907). The relative risk of 0.962 is precisely estimated (95% CI 0.952 to 0.972) and is highly statistically significantly below one. (Z = 7.480, P < 0.001).

**Table 2:** Confirmed COVID-19 cases in Spain and Italy

|  | **Ages 30-39** | **40-49** | **Forties / thirties** | **40s/(30s+40s)** |
| --- | --- | --- | --- | --- |
| Spain cases | 20,282 | 31,488 | 1.553 | 0.608 |
| Italy cases | 14,907 | 25,644 | 1.720 | 0.633 |
| Spain / Italy |  |  |  | 0.962<br>95% CI (0.952 to 0.972)<br>P < 0.001 |

*Note:* Data are for April 28, 2020 in Italy and May 1, 2020 in Spain. Almost everyone in Spain who were ages 40-49 were born while Spain had a policy of universal BCG vaccination at birth.

In Italy, which never had BCG, there are over four deaths from COVID-19 among people in their 40s for each death among people in their 30s (223 vs. 49). In Spain, in contrast, there are just under three deaths among people in their 40s for each death of people in their 30s (22 vs. 64). While this difference in ratios is large, few people in these cohorts died. Thus, the relative risk of 0.930 has a wide 95% confidence interval (0.849 to 1.017) and is not statistically significant from unity at conventional levels (P = 0.11).

### Adjusting for different age and sex compositions

The above calculations examine the counts of illness, without accounting for difference in age structures across the nations.

Older people are more vulnerable to COVID. Thus, a potential source of bias arises if one nation had a higher share of their population in their 40s (not their 30s). The age and sex structures of the two nations are almost identical at the decade-to-decade level (Table 3). Both nations had almost about 29% more people in their 40s than in their 30s. In addition, both nations also have very close to 50% male in these age groups. This similarity implies the above analysis is not biased by one nation having a larger share of the population in the higher-risk age decades.^18^

**Table 3:** Mortality from COVID-19 counts in Spain and Italy

|  | Age 30-39 | 40-49 | 40s/(30s+40s) |
| --- | --- | --- | --- |
| Spain deaths | 22 | 64 | 0.763 |
| Italy deaths | 49 | 223 | 0.821 |
| Relative risk (Spain / Italy) |  |  | 0.929<br>CI (0.850 to 1.01)<br>P = 0.1095 |

Note: Data are from April 28, 2020 in Italy and May 1, 2020 in Spain.

The two nations’ age structures differ *within* each decade-long cohort (Table 3). In Italy, 47% were the younger half of each decade (that is, ages 30–34 or 40–44). In Spain, a slightly lower share of the cohort in their 30s is in the younger half of their thirties (44%), while slightly more of the cohort in their 40s were in the younger half of their forties (51%). Given that mortality from COVID-19 rises strongly with age, the higher share of relatively younger 40s in Spanish national data will reduce that cohort’s COVID death rate.

**Table 4:** Share of each nation’s population by half decade of age and by gender (national data)

|  | Spain |  | Italy |  |
| --- | --- | --- | --- | --- |
|  | Men | Women | Men | Women |
| 30-34 | 2.9 | 2.9 | 2.8 | 2.8 |
| 35-39 | 3.7 | 3.7 | 3.1 | 3.1 |
| 40-44 | 4.4 | 4.3 | 3.6 | 3.6 |
| 45-49 | 4.2 | 4.1 | 4.0 | 4.1 |
| Share in 40s / share in 30s | 1.30 | 1.27 | 1.29 | 1.31 |
| Share in 40s / share in 30s (men + women) | 1.29 |  | 1.30 |  |

Source: Populationpyramid.net, 2019 data, accessed May 2, 2020

When we do the age adjustment (described above) for rates of confirmed cases, the relative risk is 0.967. This estimate is almost identical to the unadjusted estimate, 0.964.

This calculation made the strong assumption that the growth rate in outcome ratios decade-to-decade in Italy is also the growth rate across each half-decade in each nation. It turns out this estimate is not sensitive to different growth rates over a decade. Reassuringly, the results similar if we use the implied growth rates comparing cases and mortality of Italians in their 50s versus those in their 40s. At the same time, these alternative estimates still assume the growth rate across half-decades within a decade is the same for each nation and decade.

## Discussion

### Key results

The Spanish cohort that received BCG had a relative risk of 0.962 of having a confirmed case of COVID-19 (95% CI 0.952 to 0.972, P < 0.001). This cohort also had lower mortality risk (relative risk 0.929), though the reduction was not statistically significant CI 0.850 to 1.01, P =0.11).

### Limitations

Interpretation of the effect sizes is easiest if no Spaniards in their thirties and no Italians received BCG vaccination, while 100% or Spaniards in their forties did. Those assumptions do not hold. For example, migrants received BCG based on their childhood location. Spain never reached 100% coverage of BCG. The 1981 stopping date of BCG in Spain implies some people in their late 30s in February through April 2020 received BCG as children, but are part of the “thirties” (non-BCG) cohort in this analysis. As noted above, Catalonia and the Basque Region ended their BCG programs at different years than the national program (1974 and 2013, respectively). In addition, Italy and post-1981 Spain never strove for 0% BCG coverage. High-risk groups were immunized in all of Italy, and some Italian regions had protocols for widespread BCG. This fuzziness will tend to bias down the estimated effects of BCG on COVID outcomes.

The interpretation of the gap as causal is biased down because lower infections among those in their 40s will help protect those in the younger age group. That is, there are positive spillovers from the treatment group (those with BCG immunization) to the control group (nearby people without BCG).^19^

In addition, the ten-year window for comparison groups leaves room for omitted factors to affect the outcome. For example, we were able to do a coarse adjustment for differences in the age distribution. In addition, the relationship of risk factors such as heart disease and obesity with age may not be the same in the two nations.

### Interpretation

These findings provide a shred of evidence that BCG vaccination helps protect against COVID-19. The analysis uses very coarse measures of exposure to BCG vaccination. Data limitations also required use of broad age ranges and no covariates other than a coarse adjustment for age distributions.

In short, an important reason of releasing this analysis is to encourage analysts with access to more detailed data to perform a regression discontinuity study with a narrower bandwidth. If we had data with outcomes by age or year of birth, we could redo the analysis on confirmed cases with a narrower bandwidth of ages; for example, comparing ages 36–38 to those ages 40–42. As the age window narrows, the study design becomes closer to a regression discontinuity design, which is more credible.^20^

More specific age data would permit an analysis of the effect of Spain introducing universal BCG at birth in 1965. The larger sample size would permit more precision on estimating how exposure to a BCG program affects mortality.

Precision would increase further if the analysis used geographic data on BCG programs and patients. If sample sizes permitted, one could also examine the discontinuities of region-specific BCG programs. For example, Catalonia ended BCG prior to the rest of Spain (1970 versus 1981) and now has an above-average rate of COVID-19.^21^ The Basque region continued BCG until 2013 and has a slightly below-average rate of COVID-19. It would be interesting to see if those in their 40s who were born in Catalonia have a higher infection rate than those born in the rest of Spain, while those in their 30s have lower rates in the Basque region. As noted above, southern regions in Italy had high tuberculosis in the 1960s.^22^ It is likely those regions had higher rates of BCG immunization. In contrast, COVID-19 has almost entirely affected northern Italy. Better data on Italy’s regional BCG programs would permit additional comparisons.

It is also reasonable to look for other nations to use as comparisons (e.g., France and Portugal). It would also be useful to have data on additional outcomes such as hospitalizations.^23^

## Data Availability

I can share the data I downloaded on request.

## Footnotes

1 Haas School of Business, University of California, Berkeley. I have no conflicts of interest.

2 Higgins, J. P. et al. Association of BCG, DTP, and measles containing vaccines with childhood mortality: systematic review. *BMJ* 355, i5170 (2016). Hollm-Delgado, M. G., Stuart, E. A. & Black, R. E. Acute lower respiratory infection among Bacille Calmette-Guerin (BCG)-vaccinated children. *Pediatrics* 133, e73–e81 (2014).

3 Zwerling, Alice, Marcel A. Behr, Aman Verma, Timothy F. Brewer, Dick Menzies, and Madhukar Pai. “The BCG World Atlas: a database of global BCG vaccination policies and practices.” *PLoS medicine* 8, no. 3 (2011). https://www.ncbi.nlm.nih.gov/pmc/articles/PMC3062527/

4 Hegarty, Paul K., Ashish M. Kamat, Helen Zafirakis, and Andrew Dinardo. “BCG vaccination may be protective against Covid-19.” *preprint* (March 2020). DOI: 10.13140/RG.2.2.35948.10880 Sam Ebenezer Athikarisamy. “Does BCG bolster one’s immunity against COVID-19?” Letter to the editor, *BMJ*, 20 March 2020. https://www.bmj.com/content/368/bmj.m1252/rr-4 Masayuki Miyasaka. “Is BCG vaccination causally related to reduced COVID-19 mortality?”Commentary, *EMBO Molecular Medicine*, (2020) https://doi.org/10.15252/emmm.202012661 Miller, Aaron, Mac Josh Reandelar, Kimberly Fasciglione, Violeta Roumenova, Yan Li, and Gonzalo H. Otazu. “Correlation between universal BCG vaccination policy and reduced morbidity and mortality for COVID-19: an epidemiological study.” *medRxiv* (2020). https://www.medrxiv.org/content/10.1101/2020.03.24.20042937v1

5 Roni Caryn Rabin. “Can an Old Vaccine Stop the New Coronavirus?” *New York Times*. April 3, 2020 Updated April 5, 2020. https://www.nytimes.com/2020/04/03/health/coronavirus-bcg-vaccine.html

6 Hensel, Janine, Daniel J. McGrail, Kathleen M. McAndrews, Dara Dowlatshahi, Valerie S. LeBleu, and Raghu Kalluri. “Exercising caution in correlating COVID-19 incidence and mortality rates with BCG vaccination policies due to variable rates of SARS CoV-2 testing.” *medRxiv* (2020).

7 Guan WJ et al. Comorbidity and its impact on 1590 patients with Covid-19 in China: A Nationwide Analysis. *Eur Respir J* 2020 Mar 26; [e-pub]. (https://doi.org/10.1183/13993003.00547-2020)

8 Anzolin, Elisa; Amante, Angelo (21 February 2020). “Coronavirus outbreak grows in northern Italy, 16 cases reported in one day”. Thomson Reuters. Archived from the original on 21 February 2020. Retrieved 21 February 2020. Ansede, Manuel (2020–04–22). “El análisis genético sugiere que el coronavirus ya circulaba por España a mediados de febrero”. *El Pa País* (in Spanish). Retrieved 2020–04–23.

9 “Coronavirus latest: Britain’s Prince Charles tests positive for Covid-19”*. South China Morning Post. 2020–03–25. Retrieved 2020–03–25*. The United States later passed both Italy and Spain in the number of confirmed COVID-19 cases.

10 https://www.worldometers.info/coronavirus/, accessed May 11, 2020.

11 Higgins, J. P. et al. Association of BCG, DTP, and measles containing vaccines with childhood mortality: systematic review. *BMJ* 355, i5170 (2016). Hollm-Delgado, M. G., Stuart, E. A. & Black, R. E. Acute lower respiratory infection among Bacille Calmette-Guerin (BCG)-vaccinated children. *Pediatrics* 133, e73–e81 (2014).

12 van der Meer, Jos WM, Leo AB Joosten, Niels Riksen, and Mihai G. Netea. “Trained immunity: a smart way to enhance innate immune defence.” *Molecular immunology* 68, no. 1 (2015): 40–44. Blok, Bastiaan A., Rob JW Arts, Reinout van Crevel, Christine Stabell Benn, and Mihai G. Netea. “Trained innate immunity as underlying mechanism for the long-term, nonspecific effects of vaccines.” *Journal of leukocyte biology* 98, no. 3 (2015): 347–356.

13 Zwerling, Alice, Marcel A. Behr, Aman Verma, Timothy F. Brewer, Dick Menzies, and Madhukar Pai. “The BCG World Atlas: a database of global BCG vaccination policies and practices.” *PLoS medicine* 8, no. 3 (2011). https://www.ncbi.nlm.nih.gov/pmc/articles/PMC3062527/. Catalonia stopped universal vaccination in 1974, while the Basque Country continued until 2013. Asociacion Espanola de Vacunologia, “Vacunas disponibles.” 20 Dec. 2018. https://www.vacunas.org/vacunasdisponibles-tuberculosis/ Some Italian regions had programs for widespread BCG for some of the 1965–1981 period, but I am unable to determine which regions. During the early 1960s rates of tuberculosis were highest in the south of the nation. (L’eltore, G. “Tuberculosis. in Italy Today.” *Lotta contro la Tubercolosi* 34, no. 3/4 (1964): 297–309.) Thus, it is likely that most immunizations were in the south. In contrast, COVID cases are concentrated in northern Italy. Thus, it is likely that regional BCG immunization programs in Italy were not relevant for many people in the regions where most COVID cases occurred.

14 Centro de Coordinación de Alertas y Emergencias Sanitarias, “Actualización nº 92. Enfermedad por el coronavirus (COVID-19). 01.05.2020 (datos consolidados a las 21:00 horas del 30.04.2020)” https://www.mscbs.gob.es/profesionales/saludPublica/ccayes/alertasActual/nCov- China/documentos/Actualizacion_92_COVID-19.pdf accessed May 1, 2020.

15 “Characteristics of SARS-CoV-2 patients dying in Italy Report based on available data on April 28th,” 2020. https://www.epicentro.iss.it/coronavirus/bollettino/Bollettino-sorveglianza-integrata-COVID-19_28-aprile-2020.pdf

16 Lisa Schnirring. “As Italy COVID-19 cases soar, WHO tackles PPE, test shortages.” CIDRAP News, Mar 20, 2020. https://www.cidrap.umn.edu/news-perspective/2020/03/italy-covid-19-cases-soar-who-tackles-ppe-test-shortages, accessed May 8, 2020.

17 Carl Heneghan, Jon Brassey, Tom Jefferson. “COVID-19: What proportion are asymptomatic? The Centre for Evidence-Based Medicine, April 6, 2020. https://www.cebm.net/covid-19/covid-19-what-proportion-areasymptomatic/, accessed May 8, 2020.

18 In Italy most cases are in Lombary, with large concentrations in Emilia-Romagna, Piedmont and Veneto. (https://www.statista.com/statistics/1099375/coronavirus-cases-by-region-in-italy/), May 7 2020). Those regions have mean ages that are fairly typical of Italy. https://www.statista.com/statistics/569187/average-age-of-thepopulation-in-italy-by-region/ In Spain cases are concentrated in Madrid and Catalonia. Again, those age distributions are similar to those of the rest of the nation.

19 Experiments related to infectious diseases usually violate the “Stable Unit Treatment Value Assumption” of no spillovers.

20 Hahn, Jinyong, Petra Todd, and Wilbert Van der Klaauw. “Identification and estimation of treatment effects with a regression discontinuity design.” *Econometrica* 69, no. 1 (2001): 201–209.

21 https://covid19.isciii.es/, last accessed May 7, 2020.

22 L’eltore, G. “Tuberculosis. in Italy Today.” *Lotta contro la Tubercolosi* 34, no. 3/4 (1964): 297–309.

23 An alternative study design is to examine whether migrants from nations with universal BCG should have lower rates of severe COVID symptoms. For example, one might compare rates of hospitalization for COVID and other severe symptoms of migrants who received BCG as children and then moved to a nation without BCG to their siblings born in the nation without BCG. More generally, comparing migrants with BCG who came here at a young age to children of migrants may be informative.

